# Associations of the Patient Safety Screener-3 With Depression and Suicide Risk: A Nationwide Cross-Sectional Study in Japan

**DOI:** 10.64898/2026.08.30.26361711

**Authors:** Kana Kiryu, Hidetaka Tamune, Keitaro Takahashi, Hirohisa Fujikawa, Hiroyuki Harada, Sho Fukui, Kazuya Nagasaki, Yuji Nishizaki, Tadafumi Kato, Yasuharu Tokuda

## Abstract

**Aim:** The Patient Safety Screener-3 (PSS-3) is a brief suicide-risk screening tool. Item 1 of this scale assesses depressive mood but is not included in the total score. We examined the association of item 1 with depressive symptom severity and characterized the suicide-related risk captured by PSS-3 total positivity.

**Methods:** We conducted a nationwide cross-sectional survey among resident physicians in Japan. Associations between PSS-3 item 1 endorsement and Patient Health Questionnaire-9 (PHQ-9) scores were evaluated using the Wilcoxon rank-sum test. Diagnostic performance of item 1 was evaluated using PHQ-9 positivity (≥10) as reference standard. We also compared Short-form Scale for Suicide Ideation (SIS-6) scores according to PSS-3 total positivity and PHQ-9 item 9 positivity.

**Results:** A total of 1,844 participants were included. PSS-3 item 1 was endorsed by 443 physicians (24.0%), and 47 (2.5%) met the criteria for PSS-3 total positivity. Item 1 showed 79.3% sensitivity and 79.5% specificity for PHQ-9 positivity. SIS-6 scores were higher in the PSS-3 total-positive group than in the total-negative group (median [IQR], 6 [5–9] vs 0 [0–1]; p<0.001). The SIS-6 showed a higher area under the receiver operating characteristic curve (AUC) and Youden index using PSS-3 total positivity (AUC, 0.961; optimal cutoff, 3) than PHQ-9 item 9 positivity (AUC, 0.907; optimal cutoff, 2).

**Discussion:** PSS-3 may support brief, simultaneous screening for depressive symptoms and suicide-related risk. Compared with PHQ-9 item 9, PSS-3 may capture a more severe spectrum of suicide-related risk. PSS-3 may facilitate identification of individuals requiring further mental health assessment.

**Graphical Abstract:** 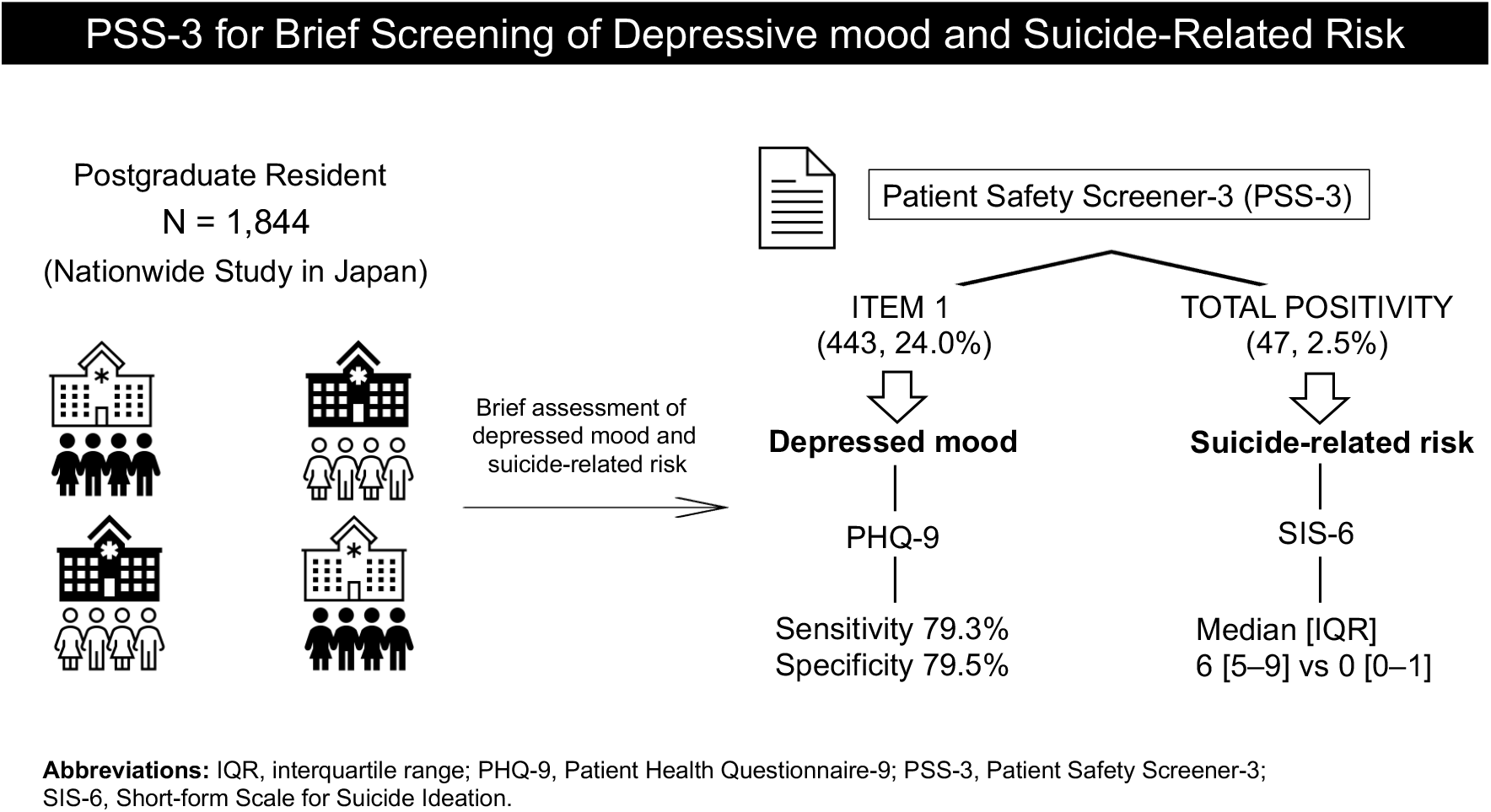

## 1. Introduction

Depression is a common and clinically important disorder that is closely associated with suicidal behavior and suicide risk (Carter et al., 2017). The diagnosis of depressive disorders is usually based on symptom criteria defined in the Diagnostic and Statistical Manual of Mental Disorders (DSM). The Patient Health Questionnaire-9 (PHQ-9), which directly reflects the nine DSM symptom criteria for depression, is one of the most widely used instruments for screening and assessing the severity of depressive symptoms (Kroenke et al., 2001). The US Preventive Services Task Force (USPSTF) recommends screening all adults for depression when appropriate systems are in place to ensure accurate diagnosis, effective treatment, and follow-up (US Preventive Services Task Force, 2023).

Suicide is also a major public health issue worldwide and a preventable cause of death. Identifying individuals who need support and linking them to appropriate assessment and intervention are central to suicide prevention (World Health Organization, 2021). Although repeated, multi-informant assessment by trained clinicians is the most reliable approach to suicide risk assessment (Calati et al., 2019), universal assessment by specialists is not feasible in real-world settings. Thus, screening for suicidal risk can help triage individuals who require additional assessment and management and may facilitate timely intervention (Stapper et al., 2026).

Evidence-based interventions for depression and suicide prevention are commonly organized within a framework that distinguishes between universal, selective, and indicated interventions (World Health Organization, 2014; Cuijpers, 2025): universal interventions target the whole population, selective interventions target high-risk groups, and indicated interventions target specific vulnerable individuals (Pirkis et al., 2024). Accordingly, we speculated that brief screening tools that can assess both depressed mood and suicide-related risk may help bridge selective and indicated interventions.

The Patient Safety Screener-3 (PSS-3) is a brief screening instrument developed to identify suicide risk. It consists of three principal items which assess depressed mood, active suicidal ideation, and lifetime suicide attempt. Item 1, which assesses depressed mood, was originally included to facilitate the transition to the subsequent suicide-related questions and is not part of the PSS-3 scoring algorithm. We previously reported that endorsement of item 1 was independently associated with a longer hospital stay, suggesting that it may capture clinically relevant characteristics (Tamune et al., 2025). We therefore considered that the PSS-3 may provide a practical instrument for assessing both depressed mood and suicide-related risk within a single brief screening process. However, the clinical performance of PSS-3 item 1 as a screening indicator of depressive symptoms has not been systematically evaluated.

In this study, we examined whether endorsement of PSS-3 item 1 was associated with depressive symptom severity and investigated the characteristics of suicidal ideation captured by PSS- 3 total positivity.

## 2. Methods

### 2.1. Study design and participants

We conducted a nationwide cross-sectional study using an anonymous, online, self-administered questionnaire. We surveyed postgraduate year (PGY) 1 and 2 resident physicians after they completed the General Medicine In-Training Examination (GM-ITE) in January 2026. The GM-ITE is a nationwide computer-based examination taken by approximately half of resident physicians in Japan and is validated for the assessment of clinical knowledge (Tamune et al., 2024; Harada et al., 2026). For the present study, only GM-ITE examinees who provided informed consent were included. Participants were excluded if they did not complete the PSS-3, which was the primary exposure measure.

### 2.2. Measures

#### 2.2.1 Depression-related symptoms

PSS-3 item 1 (“In the past two weeks, have you felt down, depressed, or hopeless?”) is intended to assess depressed mood, but does not contribute to the total score. Depressive symptoms were assessed using the Patient Health Questionnaire-9 (PHQ-9) (Muramatsu et al., 2018), a widely used and psychometrically validated instrument with established internal consistency and criterion validity across diverse populations (Kroenke, 2021). Total scores range from 0 to 27, with higher scores indicating greater depressive symptoms. PHQ-9 positivity was defined as a total score ≥10.

#### 2.2.2 Suicide ideation-related symptoms

The PSS-3 score was calculated by summing the scores of items 2, 3, and 3a, to give a score ranging from 0 to 4. PSS-3 total positivity was defined as a score of ≥2 (Tamune et al., 2024).

The Scale for Suicide Ideation (SSI) is the Japanese cultural adaptation of the Beck Scale for Suicide Ideation, a widely validated instrument that assesses the intensity, severity, and characteristics of suicidal ideation (Otsuka et al., 1998). The Short-form Scale for Suicide Ideation (SIS-6) is a six-item abbreviated version of the SSI whose reliability and validity was demonstrated in an internet-based survey of Japanese adults aged 20–69 years (Sueki et al., 2019). Scores range from 0 to 12, with higher scores indicating greater suicidal ideation.

To identify suicidal ideation, we utilized the ninth item of PHQ-9 (‘thoughts that you would be better off dead, or thoughts of hurting yourself in some way?’). This single item has been extensively used to assess suicidal ideation in previous epidemiological studies (Wang et al., 2020).

### 2.3. Data analysis

Participant characteristics were summarized using descriptive statistics. Categorical variables are presented as frequencies and percentages, and continuous variables as medians and interquartile ranges (IQR), as appropriate.

#### 2.3.1 Depression analyses

The association between PSS-3 item 1 endorsement and PHQ-9 score was examined using the Wilcoxon rank-sum test. Effect sizes for the Wilcoxon rank-sum tests were expressed as the rank-biserial correlation with 95% confidence intervals (CIs).

The association between PSS-3 item 1 endorsement and PHQ-9 positivity was examined using Pearson’s χ² test. Using the PHQ-9 as the reference standard for clinically significant depressive symptoms, we calculated the sensitivity and specificity, positive and negative predictive values, and receiver operating characteristic (ROC) analysis for PSS-3 item 1 endorsement. PHQ-9 item 9 endorsement was defined as any response other than “not at all” (score ≥ 1).

#### 2.3.2 Suicide-related analyses

Because the SIS-6 does not have an established cutoff score, SIS-6 scores were compared between the PSS-3 total-positive and -negative groups using the Wilcoxon rank-sum test. Effect sizes for the Wilcoxon rank-sum tests were expressed as the rank-biserial correlation with 95% confidence intervals.

As a supplementary analysis, the association between the PSS-3 score and SIS-6 score was assessed using Spearman’s rank correlation. In addition, to compare the characteristics of suicidal ideation captured by the PSS-3 and PHQ-9 item 9, ROC analyses were conducted using the SIS-6. PSS- 3 total positivity and PHQ-9 item 9 endorsement were used separately as reference standards, and the area under the curve (AUC) and Youden index were calculated for each analysis.

### 2.4. Missing data and statistical software

Responses of “Patient unable to complete” and “Patient refused” were excluded from analyses in which PSS-3 item 1 served as the study variable. For analyses using the PSS-3 score, these responses were assigned a score of 0 per the original PSS-3 scoring algorithm. For each analysis, only participants with complete data for all variables included in that specific analysis were retained (complete-case analysis). Statistical significance was assessed using two-tailed tests, with p values < 0.05 considered to indicate statistical significance. All analyses were conducted using R version 4.5.2.

### 2.5. Ethical considerations

This study was approved by the Ethics Committee of the Japan Institute for Advancement of Medical Education Program (approval no. 25-11). Informed consent was obtained from participants using an opt-in procedure.

## 3. Results

### 3.1. Participant characteristics

A total of 9,419 individuals were invited to participate in the survey. Of these, 2,208 completed the survey and 1,905 provided informed consent. After excluding respondents with incomplete PSS-3, 1,844 participants (19.6% of all invitees) were included in the final analysis (**Figure 1**).

**Figure 1.**
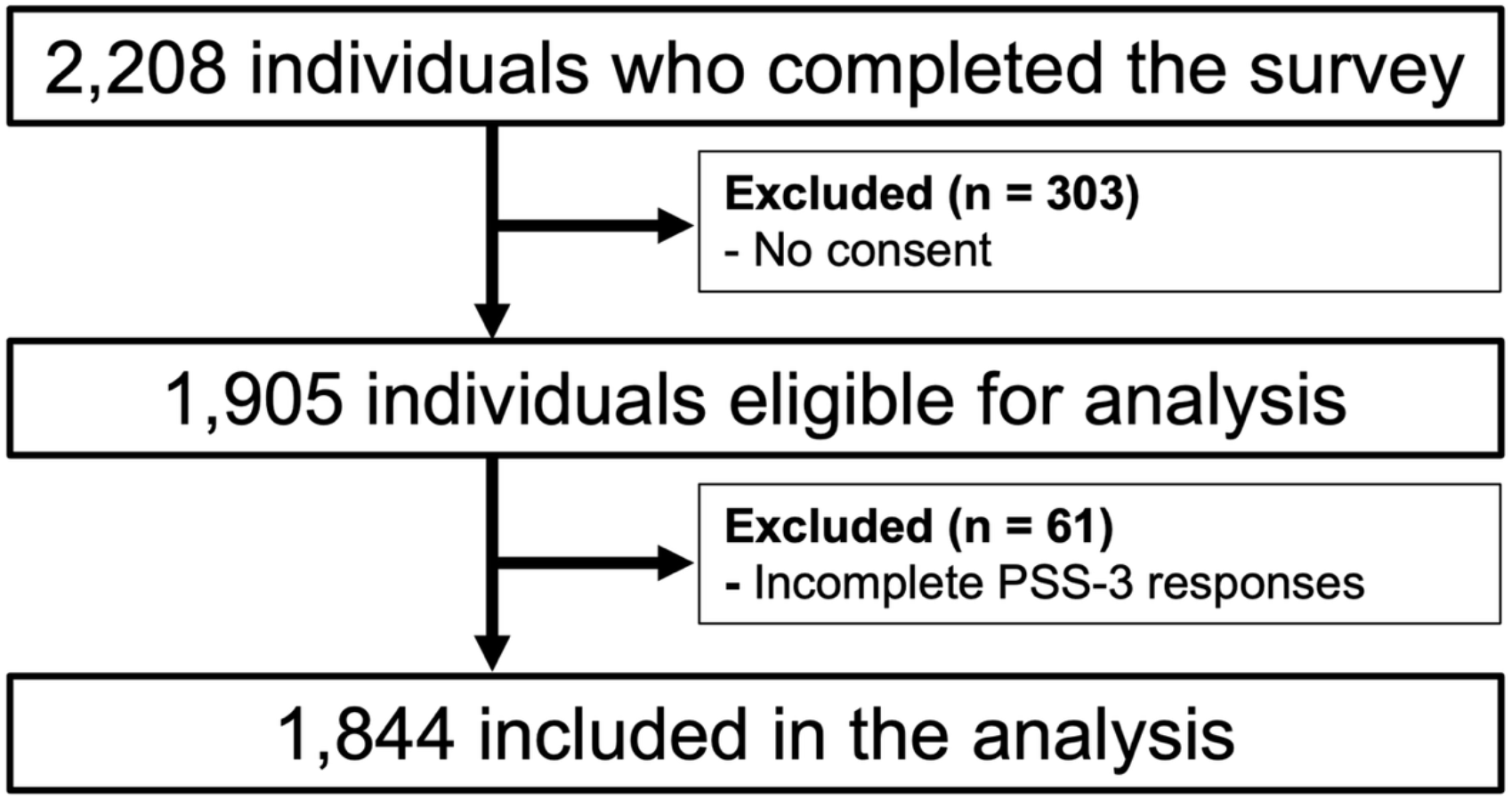
Flow diagram of study participant selection. A total of 9,419 residents completed the GM-ITE and were invited to participate in the survey. Of these, 2,208 completed the survey, and 1,905 provided informed consent and were eligible for analysis. After excluding respondents with incomplete PSS-3, 1,844 participants (19.6% of all invitees) were included in the final analysis. **Abbreviations:** GM-ITE, the General Medicine In-Training Examination; PSS-3, Patient Safety Screener-3.

**Table 1** shows participant characteristics for the overall sample stratified by PSS-3 item 1 positivity and PSS-3 total positivity. The median age of participants was 26.0 years (IQR: 26–27), and the majority were male (n = 1,173, 63.6%). Of these, 1,526 (82.8%) were at a community hospital.

**Table 1.**
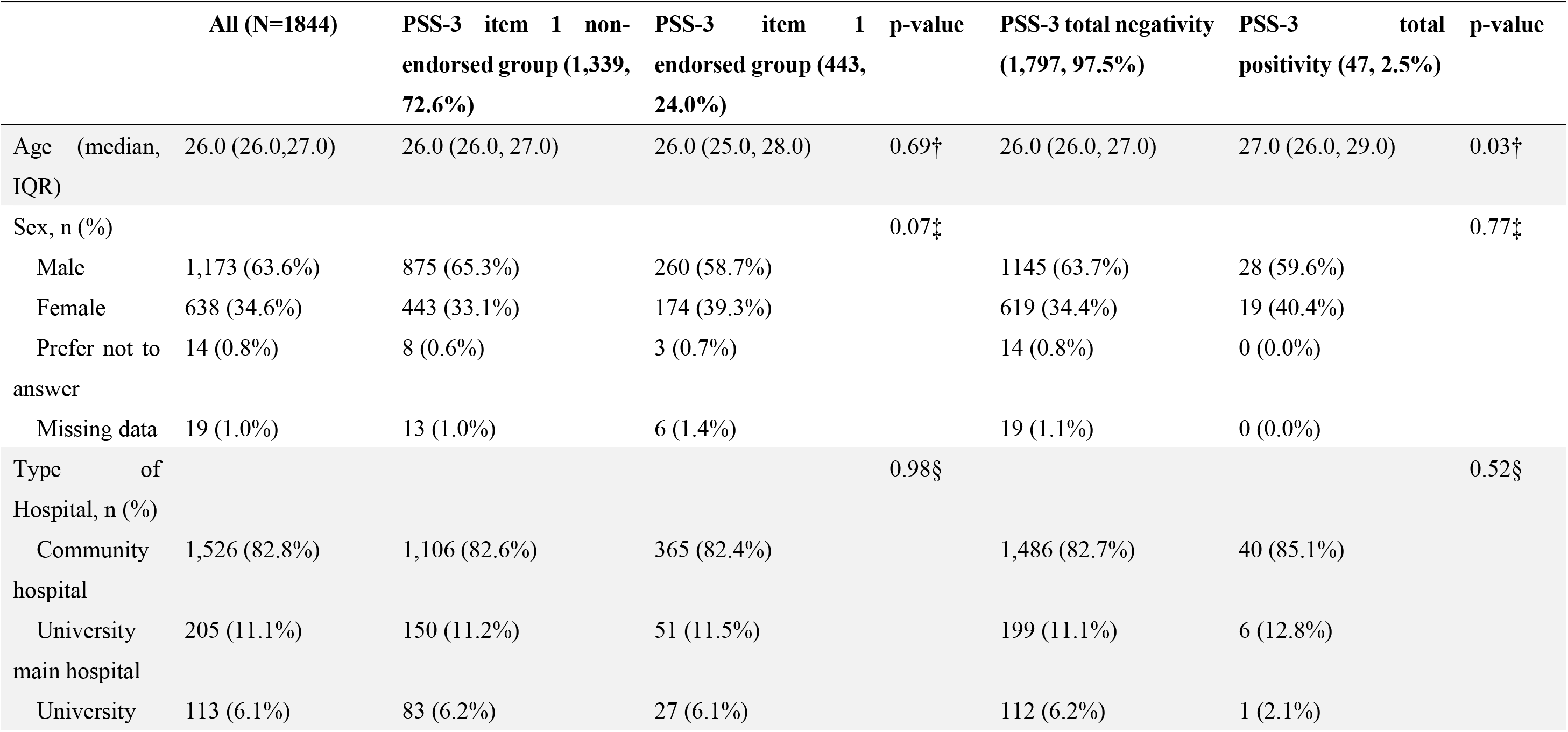
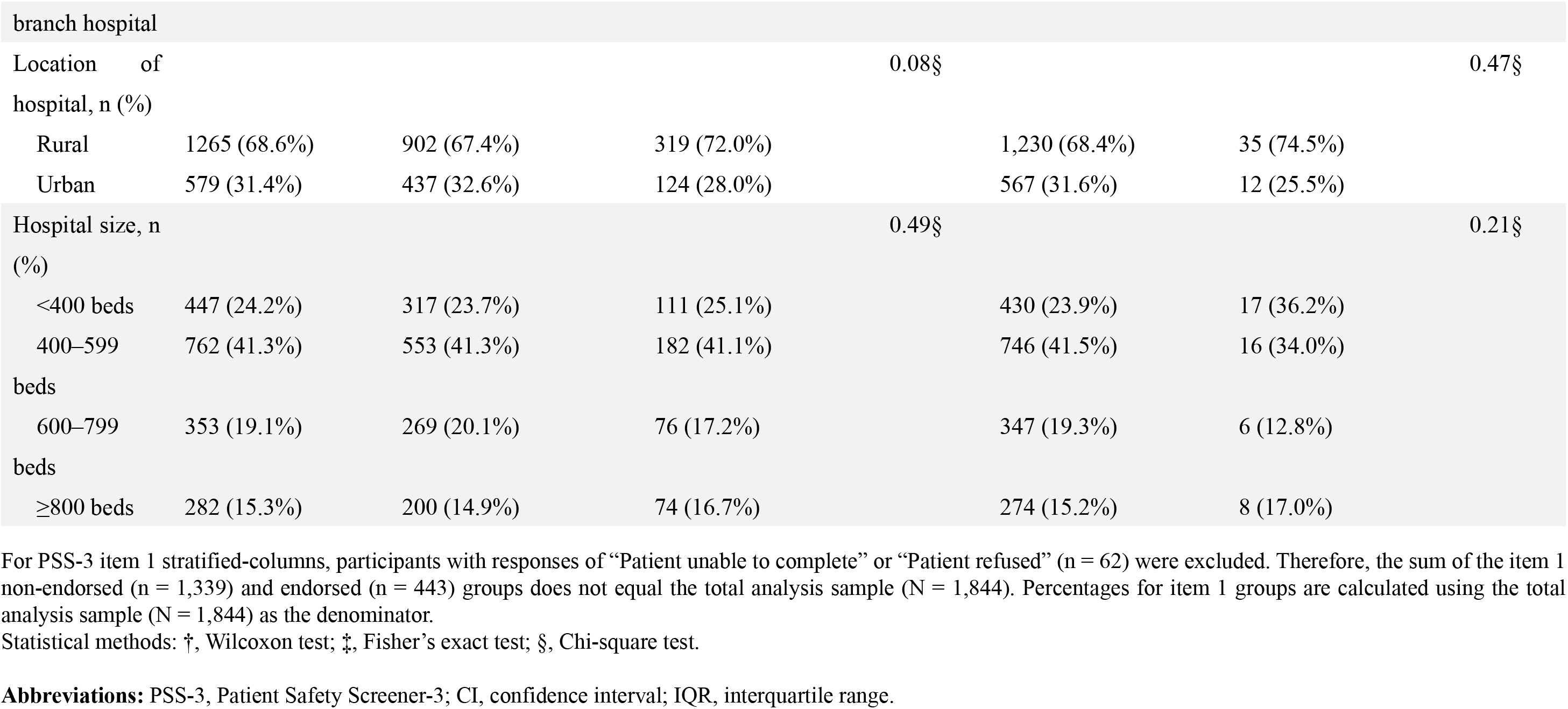
Characteristics of the participants (N = 1844)

**Figure 2** summarizes the study’s analytical framework and maps each analysis to its corresponding tables and figures in the manuscript.

**Figure 2.**
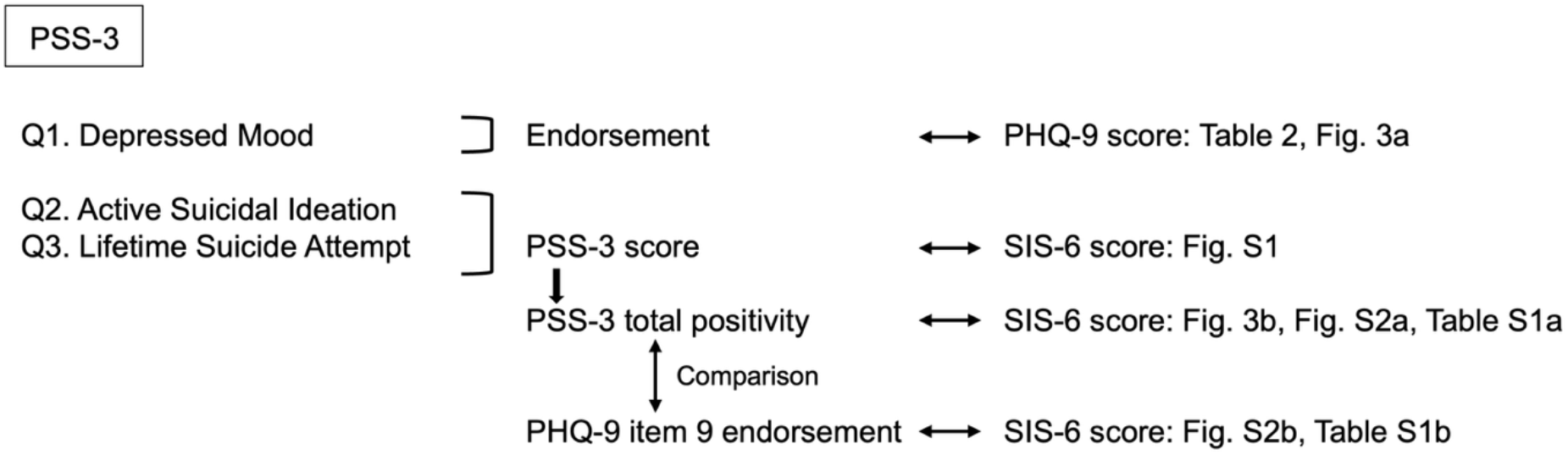
Analytical framework of the study. Overview of the analytical framework, summarizing the study variables, analytical strategy, and corresponding tables and figures. **Abbreviations:** PHQ-9, Patient Health Questionnaire-9; PSS-3, Patient Safety Screener-3; SIS-6, Short-form Scale for Suicide Ideation.

Among the 1,844 participants, 443 (24.0%) endorsed PSS-3 item 1. Participants who responded “Patient unable to complete” or “Patient refused” to item 1 (n = 62) were excluded from the item 1 stratified analyses shown in **Table 1**. Overall, 47 of 1,844 participants (2.5%) met the criteria for PSS-3 total positivity. Complete PHQ-9 data for all nine items were available for 1,741 participants, and complete SIS-6 data for 1,784 participants.

### 3.2 Depression analyses

Depression-related analyses were restricted to participants with a valid yes/no response to PSS-3 item 1 and complete responses to all nine PHQ-9 items (n = 1,684). PHQ-9 scores were significantly higher in the PSS-3 item 1 endorsed group (n = 424) than in the item 1 non-endorsed group (n = 1,260, Wilcoxon rank-sum test; median (IQR), 6 (3,10) vs. 1 (0,3), p < 0.001; rank-biserial correlation r = 0.67, 95% CI, 0.64–0.71). In addition, a significant association was observed between the PSS-3 item 1 endorsed group and the PHQ-9 positive group according to Pearson’s chi-square test (p < 0.001, **Figure 3a**).

**Figure 3a.**
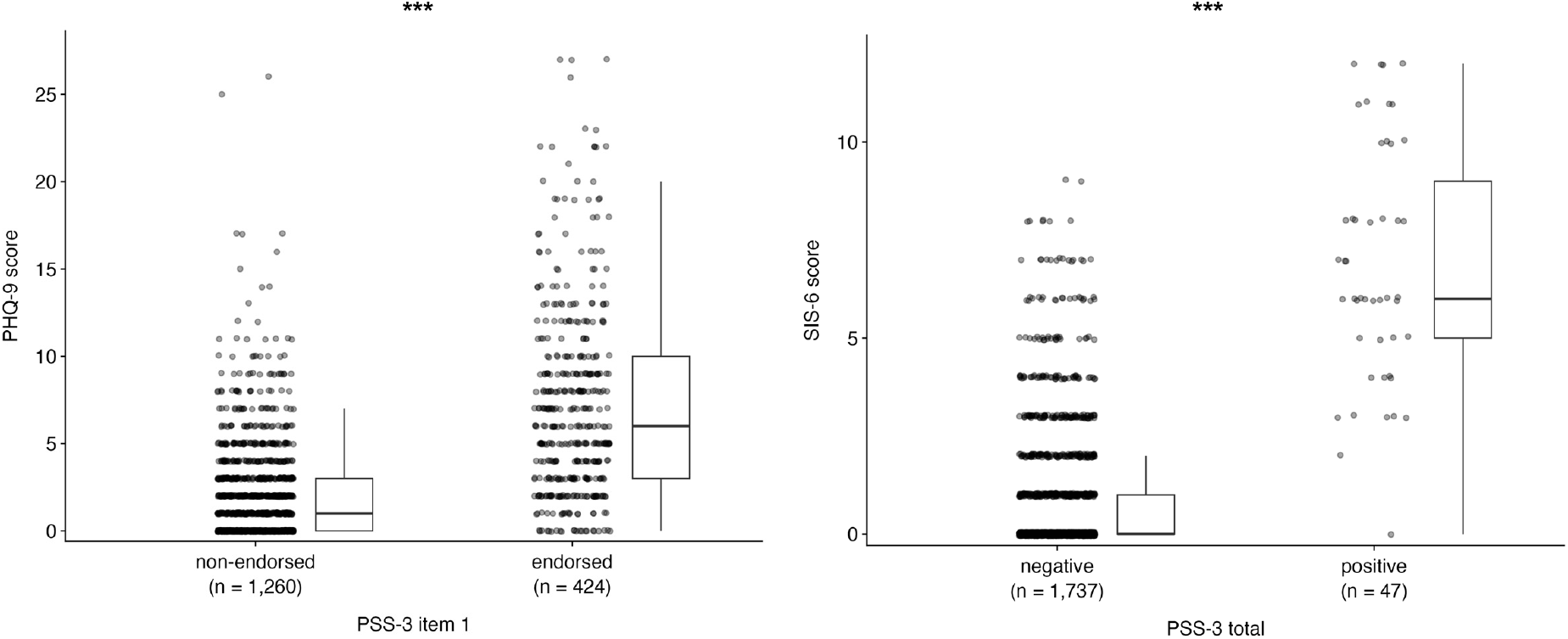
PHQ-9 scores according to PSS-3 item 1 endorsement. Figure 3b. SIS-6 scores according to PSS-3 total positivity. (a) Box-and-whisker plots with jittered individual data points showing PHQ-9 scores according to PSS-3 item 1 endorsement. Complete PHQ-9 data were available for 1,741 participants; of these, 1,684 with a valid yes/no response to PSS-3 item 1 were included in the analysis. (b) Box-and-whisker plots with jittered individual data points showing SIS-6 scores according to PSS-3 total positivity. Analyses were limited to 1,784 participants with complete scorable responses to all six SIS-6 items. **Abbreviations:** PHQ-9, Patient Health Questionnaire-9; PSS-3, Patient Safety Screener-3; SIS-6, Short-form Scale for Suicide Ideation.

Using PHQ-9 positivity as the reference standard, PSS-3 item 1 endorsement showed a sensitivity of 79.3% (71.7–85.2%), specificity of 79.5% (77.5–81.5%), and AUC of 0.794 (0.758– 0.828). The positive and negative predictive values were 25.2% (21.3–29.6%) and 97.8% (96.8–98.5%), respectively (**Table 2**).

**Table 2.** Cross-tabulation of PSS-3 item 1 endorsement and PHQ-9 positivity.

|  | PHQ-9 positivity | PHQ-9 negativity | Total |
| --- | --- | --- | --- |
| **PSS-3 item 1 endorsed** | 107 | 317 | 424 |
| **PSS-3 item 1 non-endorsed** | 28 | 1,232 | 1,260 |
| **Total** | 135 | 1,549 | 1,684 |

PHQ-9 positivity and negativity were defined as scores of $\geq 10$ and $< 10$ , respectively. PHQ-9 positivity was used as the reference standard. Sensitivity, 79.3% (107/135); specificity, 79.5% (1,232/1,549); positive predictive value, 25.2% (107/424); negative predictive value, 97.8% (1,232/1,260); AUC, 0.794 (95% CI, 0.758–0.828).
These analyses were limited to 1,684 participants with valid yes/no responses to PSS-3 item 1.
**Abbreviations:** PHQ-9, Patient Health Questionnaire-9; PSS-3, Patient Safety Screener-3.

### 3.3 Suicide-related analyses

Analyses comparing SIS-6 scores between the PSS-3 total-positive and -negative groups, as well as Spearman’s correlation and ROC analyses using PSS-3 total positivity as the reference standard, were limited to participants with complete scorable SIS-6 data (n = 1,784). SIS-6 scores were significantly higher in the PSS-3 total-positive group (n = 47) than in the -negative group (Wilcoxon rank-sum test; median (IQR), 6 (5–9) vs. 0 (0–1); p < 0.001, rank-biserial correlation r = 0.92, 95% CI, 0.89–0.94) (**Figure 3b**).

Spearman’s rank correlation analysis demonstrated a significant positive correlation between the PSS-3 score and SIS-6 score (Spearman’s ρ = 0.392, p < 0.001, **Supplementary Figure 1)**.

When PSS-3 total positivity was used as the reference standard, analyses were limited to participants with complete scorable SIS-6 data (n = 1,784). AUC for SIS-6 was 0.961 (0.926–0.983) and optimal cutoff based on the Youden index was 3 (**Supplementary Figure 2a**; **Supplementary Table 1a**). ROC analysis using PHQ-9 item 9 endorsement (n = 141 of 1,775, 7.9%) as the reference standard was restricted to participants with complete SIS-6 data and a non-missing PHQ-9 item 9 response. In this analysis, the AUC was 0.907 (0.878–0.933) and the optimal cutoff was 2 (**Supplementary Figure 2b**; **Supplementary Table 1b**).

## 4. Discussion

Endorsement of PSS-3 item 1 was associated with substantially greater depressive symptom severity among Japanese resident physicians. Using PHQ-9 positivity as the reference, endorsement of PSS-3 item 1 showed a sensitivity of 79.3% and specificity of 79.5% for depression. In addition, participants with PSS-3 total positivity had markedly higher SIS-6 scores than those without total positivity. Collectively, these findings suggest that the PSS-3 may provide useful information on depressive symptoms and suicide-related risk within a single brief screening process.

The PSS-3 was originally developed as a suicide risk screening tool. Item 1 was included to facilitate the transition to potentially sensitive suicide-related questions rather than to contribute to the score (Boudreaux et al., 2015). The present findings extend our previous observation that endorsement of item 1 was independently associated with a longer hospital stay (Tamune et al., 2025) by showing that it is also associated with depressive symptom severity.

PSS-3 total positivity was also associated with suicidal ideation severity, as reflected by higher SIS-6 scores. This finding is consistent with previous studies showing correlations between PSS-3 and the Beck Scale for Suicide Ideation and further supports the convergent validity of the suicide-related risk component of the PSS-3 (Boudreaux et al., 2015).

In the supplementary analysis, the SIS-6 showed a numerically higher AUC and Youden index for discriminating PSS-3 total positivity than for discriminating PHQ-9 item 9 endorsement. These findings suggest that SIS-6 scores correspond more closely to the construct captured by PSS-3 total positivity than to that captured by PHQ-9 item 9 endorsement. PHQ-9 item 9 assesses passive suicidal ideation or self-harm thoughts during the preceding 2 weeks, whereas PSS-3 total positivity incorporates active thoughts of killing oneself during the preceding 2 weeks, a suicide attempt within the preceding 6 months, or both. This difference may reflect the distinction between passive and active suicidal thoughts, as well as the additional clinical information provided by recent suicide attempt history. The inclusion of recent suicide attempt history, together with the distinction between passive and active suicidal thoughts, may aid in identifying individuals with more acute suicide-related risk.

The potential value of such a brief screening tool may be greatest in populations at elevated risk of mental health problems. The mental health of resident physicians is an important global public health concern (Harvey et al., 2021), and approximately one-third of resident physicians experience depressive symptoms (Mata et al., 2015). Consistent with this concern, we previously reported that long duty hours among resident physicians were associated with depression, burnout, and high job stress (Nagasaki et al., 2022). Addressing workload and other workplace determinants is therefore a central component of prevention. The Japanese Stress Check Program provides a workplace-level approach by assessing occupational stressors, stress responses, and social support, but does not directly assess depressive symptoms or suicide-related risk. The PSS-3 may complement such workplace-level measures by providing a brief individual-level screen for depressed mood and suicide-related risk. By identifying individuals who require further assessment, the PSS-3 may help bridge selective and indicated prevention approaches.

## 5. Limitations

Our study has several limitations. First, the study population consisted exclusively of resident physicians, which may limit the generalizability of the findings. Although the PSS-3 was originally developed for use in emergency department settings, resident physicians experience substantial occupational stress and are at increased risk of depression and suicidal ideation. The findings that screening for suicidal ideation may be particularly valuable in populations with established risk factors (Stapper et al., 2026) suggests that this population remains clinically relevant. Nevertheless, further studies in more diverse populations are warranted.

Second, the survey was conducted in January. Depressive symptoms among resident physicians vary seasonally, particularly during the early months of postgraduate training, which begins in spring in Japan (Kim et al., 2024). Accordingly, the prevalence of depressive symptoms and suicidal ideation observed in this study may not reflect rates at other times of the year. However, because the primary objective of this study was to examine associations between screening measures rather than estimate prevalence, the impact of seasonal variation on the main findings is likely limited.

Third, participation was voluntary, and only 19.6% (1,844 out of 9,419 invitees) were included in the analysis. This raises the possibility of selection bias. Individuals experiencing severe psychological distress may have been less likely to participate or complete the survey, potentially leading to an underestimation of depressive symptoms and suicidal ideation.

Finally, relatively few participants had high PSS-3 scores, particularly scores of 3 or 4. The study may have had limited ability to evaluate the performance of the PSS-3 among individuals at the highest levels of suicide risk. Future studies should examine the utility of the PSS-3 in populations with theoretically higher suicide risk, such as patients with diagnosed psychiatric disorders or individuals receiving mental health care.

## 6. Conclusion

In conclusion, PSS-3 item 1, which directly assesses depressed mood, was associated with depressive symptom severity measured by the PHQ-9, whereas PSS-3 total positivity may focus on a more acute aspect of suicide-related risk than PHQ-9 item 9 alone. These findings support the potential utility of the PSS-3 as a brief and simultaneous screening tool for identifying individuals who may require further assessment for depressive symptoms and suicide-related risk.

## Data Availability

The raw data are not publicly available due to privacy or ethical restrictions but are available from the corresponding author upon reasonable request.

## Acknowledgments

The authors thank Mr. Juhei Matsumoto for his continuous technical assistance; Drs. Tomoyuki Kodama, Masaaki Sasaki, Daichi Sone, Kiyoshi Shikino, and Hiroyuki Kobayashi for their supportive comments; and all examinees and members of the Review Board of GM-ITE.

## 7. Author Contributions

KK and HT conceived the study with input from HF, YN, TK, and YT. KK analyzed the data and drafted the first manuscript supervised by HT. All authors discussed, proofread, and approved the final manuscript.

## 9. Ethics Approval Statement

This study was approved by the Ethics Review Board of the Japan Institute for Advancement of Medical Education Program (approval number: 25-11).

## 10. Patient Consent Statement

All participants provided informed consent.

## 11. Clinical Trial Registration

N/A

## 12. Conflict of Interest Statement

HT and SF have received an honorarium from JAMEP for preparing the GM-ITE exam. YN has received an honorarium from JAMEP as the GM-ITE project manager. YT, the director of JAMEP, has received an honorarium from JAMEP as a speaker at JAMEP lectures. The remaining authors declare no conflict of interest. HT, SF, YN, and YT were not involved in the data analysis.

## 13. Financial Disclosure

This study was supported by a grant from the Medical Education Promotion Foundation (HT) and Health, Labour and Welfare Policy Grants of Research on Region Medical (Number 24IA2016 to HT, HF, SF, KN, YN, and YT). It was partly supported by Takeda Science Foundation (HT), and JSPS KAKENHI Grant Number 24K18726 (HT), 26K18914 (HT), 26K18915 (KT), and 26K18939 (HH). This work, conducted by a Fellow of the Juntendo University COI-NEXT Center (HT), was also supported by JST Japan Grant Number JPMJPF2301.

## Abbreviations

IQR: interquartile range
PHQ-9: Patient Health Questionnaire-9
PSS-3: Patient Safety Screener-3
SIS-6: Short-form Scale for Suicide Ideation.

## Supporting Material List

Supplementary Table 1a. Diagnostic performance of SIS-6 cutoff scores for identifying PSS-3 total positivity

Supplementary Table 1b. Diagnostic performance of SIS-6 cutoff scores for identifying PHQ-9 item 9 endorsement

Supplementary Figure 1. Distribution of SIS-6 scores by PSS-3 total score.

Supplementary Figure 2a. ROC curve of SIS-6 for identifying PSS-3 total positivity.

Supplementary Figure 2b. ROC curve of SIS-6 for identifying PHQ-9 item 9 endorsement.

## Notes

### Author Declarations

The Ethics Review Board of the Japan Institute for Advancement of Medical Education Program gave ethical approval for this work (approval number: 25-11).

