## Supplementary Material for "Associations of the Patient Safety Screener-3 With Depression and Suicide Risk: A Nationwide Cross-Sectional Study in Japan"

**Supplementary Table 1a. Diagnostic performance of SIS-6 cutoff scores for identifying PSS-3 total positivity**

| Cutoff Score | Sensitivity (95% CI) | Specificity (95% CI) | Youden's index |
|---|---|---|---|
| ≥1 | 97.9% (88.9–99.6) | 69.9% (67.7–72.0) | 0.68 |
| ≥2 | 97.9% (88.9–99.6) | 82.4% (80.5–84.1) | 0.80 |
| **≥3** | **95.7% (85.8–98.8)** | **88.4% (86.8–89.8)** | **0.84** |
| ≥4 | 85.1% (72.3–92.6) | 93.4% (92.1–94.5) | 0.79 |
| ≥5 | 76.6% (62.8–86.4) | 96.2% (95.2–97.0) | 0.73 |
| ≥6 | 68.1% (53.8–79.6) | 97.6% (96.7–98.2) | 0.66 |
| ≥7 | 46.8% (33.3–60.8) | 98.7% (98.1–99.2) | 0.46 |
| ≥8 | 40.4% (27.6–54.7) | 99.5% (99.1–99.8) | 0.40 |
| ≥9 | 25.5% (15.3–39.5) | 99.9% (99.6–100.0) | 0.25 |
| ≥10 | 25.5% (15.3–39.5) | 100.0% (99.8–100.0) | 0.26 |
| ≥11 | 17.0% (8.9–30.1) | 100.0% (99.8–100.0) | 0.17 |
| ≥12 | 8.5% (3.4–19.9) | 100.0% (99.8–100.0) | 0.09 |

**Supplementary Table 1b. Diagnostic performance of SIS-6 cutoff scores for identifying PHQ-9 item 9 endorsement**

| Cutoff Score | Sensitivity (95% CI) | Specificity (95% CI) | Youden's index |
|---|---|---|---|
| ≥1 | 92.2% (86.6–95.6) | 73.4% (71.2–75.5) | 0.66 |
| **≥2** | **83.0% (75.9–88.3)** | **85.7% (84.0–87.4)** | **0.69** |
| ≥3 | 73.8% (65.9–80.3) | 91.3% (89.8–92.6) | 0.65 |
| ≥4 | 61.0% (52.8–68.7) | 95.8% (94.7–96.6) | 0.57 |
| ≥5 | 50.4% (42.2–58.5) | 98.1% (97.3–98.7) | 0.49 |
| ≥6 | 40.4% (32.7–48.7) | 99.0% (98.3–99.3) | 0.39 |
| ≥7 | 25.5% (19.1–33.3) | 99.5% (99.0–99.8) | 0.25 |
| ≥8 | 16.3% (11.1–23.3) | 99.8% (99.4–99.9) | 0.16 |
| ≥9 | 9.9% (6.0–16.0) | 100.0% (99.8–100.0) | 0.10 |
| ≥10 | 8.5% (4.9–14.3) | 100.0% (99.8–100.0) | 0.09 |
| ≥11 | 5.7% (2.9–10.8) | 100.0% (99.8–100.0) | 0.06 |
| ≥12 | 2.8% (1.1–7.1) | 100.0% (99.8–100.0) | 0.03 |

Sensitivity, specificity, and Youden index for each SIS-6 cutoff score are shown. Analyses included participants with complete scorable SIS-6 data. For the analysis using PSS-3 total positivity, 1,784 participants with complete responses to all six SIS-6 items were included. For the analysis using PHQ-9 item 9 endorsement, 1,775 participants with complete scorable SIS-6 data and a non-missing PHQ-9 item 9 response were included.

**Abbreviations:** CI, confidence interval; PHQ-9, Patient Health Questionnaire-9; PSS-3, Patient Safety Screener-3; SIS-6, Short-form Scale for Suicide Ideation.

**Supplementary Figure 1. Distribution of SIS-6 scores by PSS-3 total score.**

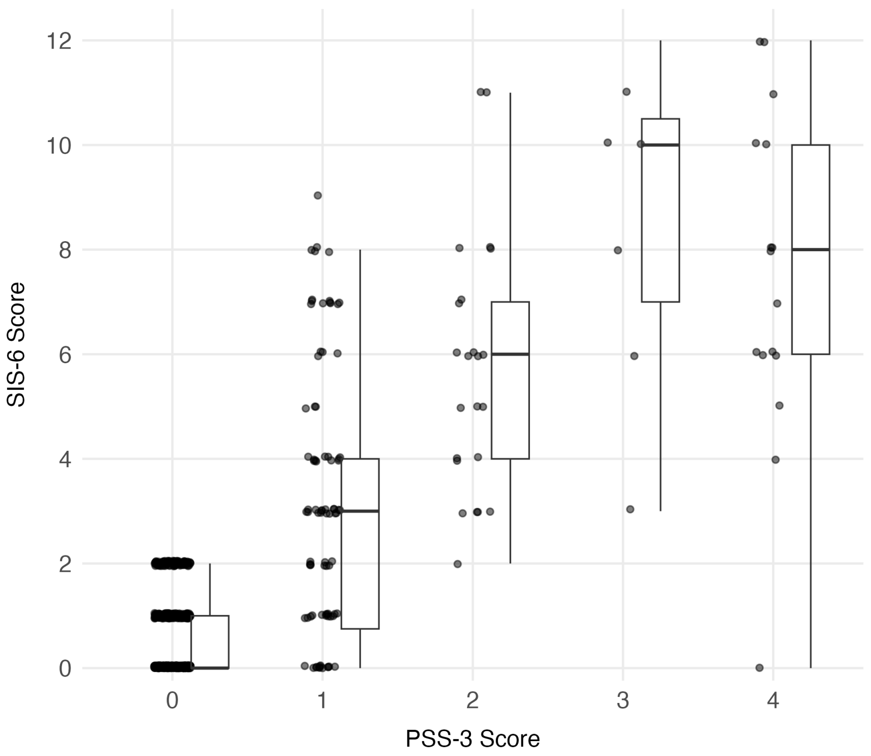


Box-and-whisker plots with jittered individual data points show SIS-6 scores by PSS-3 total score (0–4). Analysis included participants with complete scorable responses to all SIS-6 items (n = 1,784).

**Abbreviations:** PSS-3, Patient Safety Screener-3; SIS-6, Short-form Scale for Suicide Ideation.

**Supplementary Figure 2a. ROC curve of SIS-6 for identifying PSS-3 total positivity.**

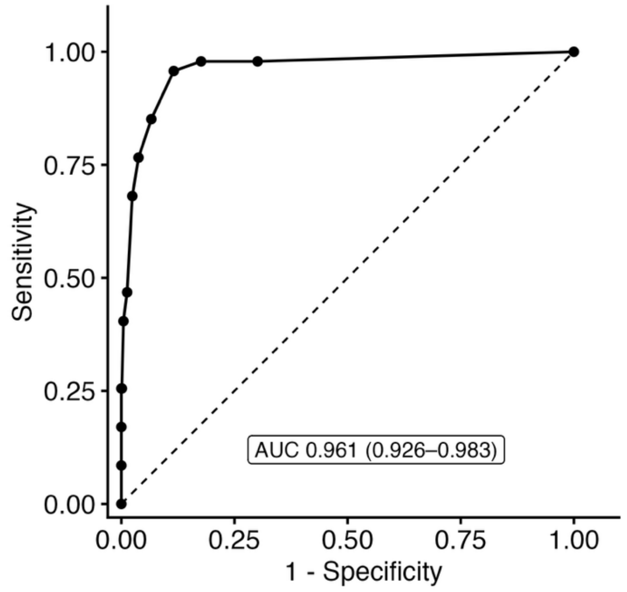


**Supplementary Figure 2b. ROC curve of SIS-6 for identifying PHQ-9 item 9 endorsement.**

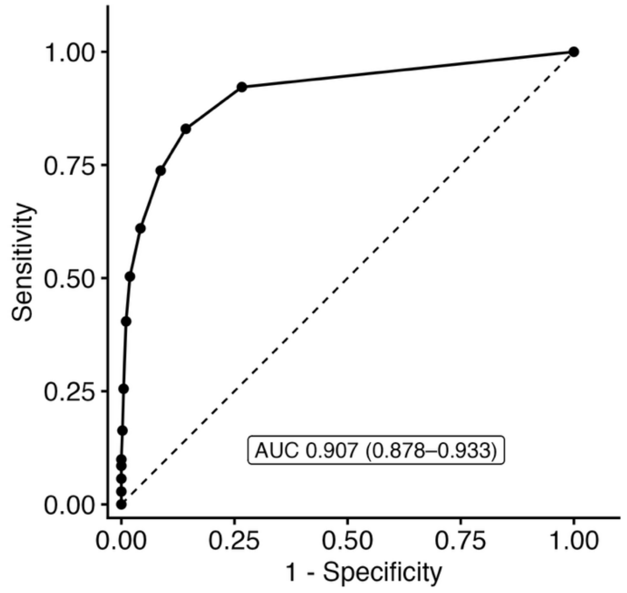


Analyses included participants with complete scorable SIS-6 data. For Supplementary Figure 2a, analyses included participants with complete scorable SIS-6 data (n = 1,784). For Supplementary Figure 2b, analyses included participants with complete scorable SIS-6 data and a non-missing PHQ-9 item 9 response (n = 1,775).

**Abbreviations:** PHQ-9, Patient Health Questionnaire-9; PSS-3, Patient Safety Screener-3; SIS-6, Short-form Scale for Suicide Ideation; ROC, receiver operating characteristic curve; AUC, area under the curve.
